# Immune evasion in prostate cancer: resolving the cold tumour paradox via a hybrid discrete-continuum computational framework

**DOI:** 10.64898/2026.03.23.26349049

**Authors:** Andile K. Ntlokwana, Edinah Mudimu, Monde Ntwasa

## Abstract

**Background:** Prostate cancer (PCa) presents a formidable clinical paradox. It is immunologically “cold” and resistant to immune checkpoint blockade (ICB), yet bulk genomic analyses consistently reveal low and non-prognostic expression of *CD274* (PD-L1), the primary molecular target of such therapies. We hypothesised that this paradox arises from a failure of current methodologies to account for two critical, interacting dimensions: the granular heterogeneity of basal gene expression (the static engine) and the spatiotemporal dynamics of adaptive resistance mediated by interferon-gamma (the adaptive engine).

**Methods:** We developed a rigorous, multi-phase computational framework integrating clinical genomics with hybrid agent-based modelling. In Phase I, we extracted and normalized *CD274* mRNA expression from the TCGA-PRAD cohort (*n* = 554) to define the empirical landscape of basal resistance. In Phase II, we developed a spatial Agent-Based Model (ABM) parameterized by this distribution to simulate clonal selection. In Phase III, we extended this into a Hybrid Discrete-Continuum model, coupling discrete agents with a reaction-diffusion Partial Differential Equation (PDE) representing the IFN-*γ* field. We simulated 50 stochastic replicates per arm across four experimental arms, including Diffusion and Induction knockouts.

**Results:** Bulk TCGA analysis confirmed low average PD-L1 expression (Median Transcripts Per Million (TPM) = 1.48; Interquartile Range (IQR): 0.91–2.14) with no prognostic value (Hazard Ratio (HR) = 1.15; 95% Confidence Interval (CI): 0.67–1.97; log-rank *p* = 0.605). However, the static ABM revealed that rare, high-expressing genomic outliers (*>* 9.0 TPM) drive persistence through Darwinian immunoediting, enriching the surviving population’s resistance by 3.86-fold. The hybrid adaptive model demonstrated a far superior survival strategy: the IFN-*γ*/PD-L1 feedback loop facilitated the emergence of “protective sanctuaries”—localised regions of high resistance at the tumour-immune interface. This mechanism increased final tumour burden by ∼4.5-fold compared to static selection alone (*p <* 0.001). Spatiotemporal analysis confirmed that resistance is not a fixed trait but a dynamic state induced by immune pressure. Diffusion knockout (*D* = 0) abolished sanctuary formation, reducing final burden by 65% (*p <* 0.001), while induction knockout (*P*_max_ = 0) reverted to static outcomes.

**Conclusions:** This study resolves the “cold” tumour paradox by demonstrating that PCa resistance is driven by a *twin engine* of rare genomic outliers and adaptive spatial dynamics. The failure of biomarkers in PCa is due to their inability to capture the dynamic mirage of adaptive sanctuaries. Our validated framework offers a platform for testing synchronised therapeutic disruptions targeting both the static genomic landscape and the dynamic cytokine signalling axis.

## **1** Introduction

### 1.1 The global burden of prostate cancer and the immunotherapy challenge

Prostate cancer (PCa) remains a leading cause of cancer-related mortality in men, with an estimated 1.4 million new cases and 375,000 deaths annually worldwide [1]. While localised disease is often curable through surgery or radiotherapy, metastatic castration-resistant prostate cancer (mCRPC) represents a lethal phenotype with a median survival of less than three years under standard therapies [1, 2]. The advent of immune checkpoint blockade (ICB), specifically antibodies targeting Programmed Death-1 (PD-1) or its ligand (PD-L1), has revolutionised the treatment of traditionally “hot” malignancies such as melanoma and non-small cell lung cancer [3, 4]. However, Phase III clinical trials (e.g., KEYNOTE-199 [5], CheckMate-650 [6]) have consistently demonstrated that PCa remains stubbornly refractory to ICB monotherapy, with objective response rates below 5% [2, 7].

### 1.2 The immunological “cold” tumour paradox

PCa is widely classified as an immunologically “cold” tumour, characterised by: (i) low infiltration of Cytotoxic T Lymphocytes (CTLs); (ii) a suppressive Tumour Microenvironment (TME) dominated by Myeloid-Derived Suppressor Cells (MDSCs), Regulatory T cells (Tregs), and M2-polarised macrophages; (iii) low tumour mutational burden (TMB); and (iv) paucity of neoantigens [2, 8, 9].

A central mechanism of immune evasion in “hot” tumours is the upregulation of PD-L1 on tumour cells, which binds to PD-1 on activated T-cells to induce exhaustion and apoptosis [10,11]. Logically, one would expect PD-L1 expression to be a key determinant of PCa outcomes. Yet a profound paradox persists:

**1. Clinical reality:** PCa effectively evades the immune system, and patients rarely respond to PD-1/PD-L1 blockade.
**2. Genomic reality:** Bulk transcriptomic analyses from large cohorts, including The Cancer Genome Atlas (TCGA) and the Stand Up To Cancer–Prostate Cancer Foundation (SU2C-PCF), consistently demonstrate that *CD274* (PD-L1) mRNA expression is low in primary PCa and lacks any significant association with biochemical recurrence, metastasis-free survival, or overall survival [12–14].

This discrepancy suggests that our current understanding, based largely on bulk averages and static single-core biopsies, is fundamentally incomplete [15, 16]. The tumour is evading the immune system, but the putative biomarker is silent.

### 1.3 Two missing dimensions: outliers and adaptive dynamics

Bulk RNA sequencing averages gene expression across millions of cells, mathematically drowning rare subpopulations of high-expressing cells that may be the functional drivers of resistance [17,18]. We hypothesise that these rare genomic outliers constitute the *first engine* of resistance: a reservoir of clones with intrinsically high basal PD-L1 expression pre-adapted to survive initial immune surveillance [19, 20].

Beyond heterogeneity, PD-L1 is not a fixed trait. Binding of Interferon-gamma (IFN-*γ*) to its receptor (IFNGR1/2) on tumour cells triggers the JAK–STAT1–IRF1 signalling cascade, leading to transcriptional upregulation of *CD274* and increased surface PD-L1 [23, 25]. This *adaptive immune resistance* creates a potent negative feedback loop: the anti-tumour response paradoxically triggers the very mechanism that suppresses it [9, 26]. Critically, IFN-*γ* spreads only a few cell diameters from its source within solid tumours (∼30–40 *µ*m), creating spatially confined signalling niches rather than uniform gradients—a biophysical constraint that drives the heterogeneous sanctuary formation we model [27]. Capturing this spatiotemporal feedback requires a Hybrid Discrete-Continuum framework coupling an ABM with reaction-diffusion PDEs [21, 28, 29].

### 1.4 Study objectives

This study tests a unified hypothesis: that PCa immune evasion is driven by a *twin engine* comprising (1) a static engine of rare genomic outliers selected by Darwinian immunoediting, and (2) an adaptive engine of IFN-*γ*-mediated phenotypic plasticity generating spatially organised protective sanctuaries. We test this hypothesis through a sequential, empirically-grounded pipeline: Phase I quantifies the heterogeneous genomic landscape of *CD274* in TCGA-PRAD (*n* = 554); Phase II links this landscape to cellular behaviour via a spatial ABM; Phase III extends the ABM into a hybrid discrete-continuum framework with explicit IFN-*γ* reaction-diffusion dynamics; and Phase IV validates each model component through mechanistic knockout controls. Together, these phases provide a computational resolution to the cold tumour paradox and identify rational targets for synchronised therapeutic disruption.

## 2 Literature review: theoretical and experimental context

### 2.1 Prostate cancer tumour microenvironment and immune evasion

The PCa TME is a complex ecosystem characterised by profound immunosuppression. Histological and transcriptomic analyses have consistently revealed: (i) low densities of CD8^+^ CTLs relative to other solid tumours; (ii) high infiltration of immunosuppressive populations including FoxP3^+^ Tregs, CD33^+^ MDSCs, and CD163^+^ M2-polarised tumour-associated macrophages (TAMs); (iii) secretion of inhibitory cytokines including TGF-*β*, IL-10, and VEGF; and (iv) physical barriers including dense desmoplastic stroma and aberrant vasculature [2, 8, 9]. This cold phenotype is actively maintained through reciprocal signalling between tumour cells and stromal components—TAMs suppress CTL function directly via PD-L1 expression and indirectly through arginase production [2, 8]—and is reinforced by spatial exclusion of CTLs from tumour-rich regions [16, 30].

### 2.2 The clinical paradox of PD-L1 in prostate cancer

Despite the well-characterised role of the PD-1/PD-L1 axis in T-cell exhaustion, bulk analyses across multiple independent cohorts have failed to establish PD-L1 as a prognostic biomarker in PCa [4, 11, 15, 17]. Chandrashekar et al. (2017) reported that *CD274* mRNA is significantly lower in tumour tissue than normal prostate, with no association with Gleason score or nodal status [12, 15]. Wang et al. (2024) confirmed a non-significant hazard ratio for biochemical recurrence (HR = 1.12; 95% CI: 0.78–1.61; *p* = 0.54) in 496 TCGA-PRAD samples [13, 16].

Zheng et al. (2022) found that PD-L1 protein was downregulated in PRAD tissue relative to adjacent normal tissue; prognostic stratification required a multiCox model integrating PD-L1, TIGIT, and six immune microenvironment indicators, underscoring the context-dependence of PD-L1 as a biomarker [9, 14].

We hypothesise that bulk measurements fail to capture two essential dimensions: (1) cellular heterogeneity, where rare high-expressing clones are averaged out [18]; and (2) spatial and temporal dynamics, where PD-L1 is induced locally and transiently at the tumour-immune interface [23, 27]. This study is designed explicitly to address both dimensions.

### 2.3 Agent-based and multi-scale modelling in computational oncology

Computational models are essential for understanding complex biological systems operating across multiple spatial and temporal scales [21,29,34]. While ODE/PDE models excel at describing population-level averages, they assume well-mixed conditions and cannot capture individual heterogeneity, discrete stochastic events, or local spatial interactions [21,29]. ABMs address these limitations by simulating autonomous agents with defined behavioural rules, allowing heterogeneous traits and spatial interactions to drive emergent population-level outcomes [22, 31]. Zhang et al. (2025) demonstrated that a stochastic ABM integrating tumour cells, CTLs, helper T cells, and Tregs shows targeted therapy with immunotherapy achieves optimal tumour control [22,32]. Bull and Byrne (2023) introduced the weighted pair correlation function (wPCF) to quantify spatial and phenotypic heterogeneity in an ABM of tumour-macrophage interactions, demonstrating the three Es of cancer immunoediting and generating distinct PCF signatures from continuous phenotypic variation [30]—a methodological advance directly applicable to our continuous PD-L1 expression model.

### 2.4 Positioning this study: empirical parameterisation and mechanistic validation

Prior ABMs incorporating the PD-1/PD-L1 axis—including the multiscale model of Gong et al. (2017) [34], the glioblastoma model of Storey and Jackson (2021) [33], and the PhysiCell platform of Ghaffarizadeh et al. (2018) [28]—have made foundational contributions. Our work introduces three advances over these prior approaches (summarised in Table 1): direct parameterisation of tumour agent phenotypes from the empirical TCGA-PRAD distribution (*n* = 554); a continuous, data-derived logistic dose-response function linking PD-L1 expression to evasion probability; and explicit reaction-diffusion PDE dynamics for IFN-*γ* validated through rigorous mechanistic knockout controls. Together, these advances allow us to resolve a specific, well-defined clinical paradox in a cold tumour type, rather than generically exploring checkpoint inhibition dynamics.

**Table 1:** Comparison of modelling approaches: this study vs. prior agent-based models of PD-1/PD-L1 interactions.

| Feature | Prior ABM approaches | This study |
| --- | --- | --- |
| PD-L1 status | Often binary (on/off) or abstracted concentration | Continuous variable sampled from empirical patient data distribution (TCGA-PRAD, $n = 554$) |
| Evasion/killing rule | Typically fixed probabilities or rule-based parameters | Evasion probability dynamically calculated via logistic dose-response function of each cell's *CD274* expression |
| Primary objective | General dynamics of checkpoint inhibition or therapy response | Resolution of a specific clinical paradox (low bulk PD-L1, poor ICB response) via cellular heterogeneity and spatial dynamics |
| Tumour type | Often generic or other specific cancers | Explicitly parameterised for prostate cancer, a ‘cold’ tumour with a defined paradox |
| IFN-γ dynamics | Often omitted or abstracted | Explicit reaction-diffusion PDE with bidirectional coupling; validated via knockout controls |

## 3 Materials and methods: integrated computational framework

Our methodological workflow is a sequential pipeline moving from empirical clinical data to increasingly sophisticated mechanistic simulations, with rigorous validation at each stage (Figure 1) [21, 29, 34].

### 3.1 Data acquisition and pre-processing

Gene expression data for *CD274* (Ensembl ID: ENSG00000120217) was obtained from The Cancer Genome Atlas Prostate Adenocarcinoma (TCGA-PRAD) project. Raw STAR-Counts files were accessed via the Genomic Data Commons (GDC) Data Portal. We developed a custom R script (Appendix A) to parse the directory structure, extract TPM (Transcripts Per Million) unstranded values, and map file IDs to patient barcodes using the metadata cart [36]. TPM nor-malisation accounts for both sequencing depth and gene length, ensuring comparability across samples [36,37]. The final dataset comprised expression values for 554 primary tumour samples, forming the empirical probability density function *f*_TCGA_(*E*) for ABM parameterisation.

### 3.2 Statistical and survival analysis

The *CD274* expression distribution was characterised using descriptive statistics (median, mean, IQR, skewness). For survival analysis, patients were stratified into ‘CD274-High’ and ‘CD274-Low’ groups at the median TPM value (1.48). Biochemical recurrence-free survival (BCR-FS) was compared using Kaplan-Meier curves and the log-rank test (*p <* 0.05) [12, 13]. A univariate Cox proportional hazards model was fitted to estimate the HR and 95% CI. We acknowledge the limited sample size for recurrence events (*n* = 73 with complete follow-up) as a constraint on statistical power [12, 13].

### 3.3 Agent-based model: conceptual framework and parameterisation

We developed a spatial ABM using the Mesa framework in Python (v3.0+) [22, 29, 34, 38]. The model operates on a 2D toroidal grid Ω ⊂ Z^2^ with dimensions *L* × *W* (baseline 50 × 50). Time evolves in discrete steps *t* ∈ N.

#### 3.3.1 Justification of 2D dimensionality

A 2D grid was selected to maximise computational efficiency during the large-scale stochastic replicates (*n* = 50 per arm, 500 steps per replicate) required for statistical validation [28, 29]. This approach serves as a representative cross-section of the TME—analogous to a histological section—while enabling high-throughput sensitivity analysis. Grid-scale sensitivity analysis (Section 4.6) confirmed that normalised dynamics are robust across spatial resolutions (30 × 30, 50 × 50, 100 × 100) [30].

#### 3.3.2 TumourCell agent

Each tumour agent *i* is initialised with a state vector **S***_i_* = {*x, y, E*_basal_*, E*_total_, pos}.

- **Basal PD-L1 expression (***E***_basal_):** Assigned at *t* = 0 by sampling with replacement from *f*_TCGA_(*E*). This value is static in the Phase II model, representing a heritable clonal trait.
- Immune evasion probability:

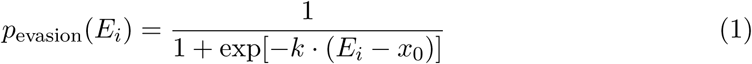

where *E_i_*is the cell’s total PD-L1 expression (TPM), *x*_0_ is the midpoint (baseline = 3.0 TPM), and *k* is the steepness coefficient (baseline = 1.0). Parameters (*x*_0_*, k*) are explored via sensitivity analysis (Section 4.6) [39–41].

- **Proliferation:** Agent *i* attempts to divide with probability *P*_prolif_ = 0.1, requiring an empty Moore neighbourhood. A daughter cell inherits identical *E*_basal_ (clonal transmission) [42].
- **Distant seeding:** If a proliferating cell finds no empty neighbours (contact inhibition), it seeds a daughter cell at a random empty location with probability *P*_seed_ = 0.001 [49,50].

#### 3.3.3 ImmuneCell agent

Immune agents represent activated CTLs performing a random walk [34, 43]:

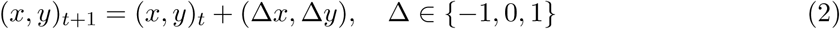

Upon co-occupation of a grid cell with a TumourCell, the probability of successful killing is:

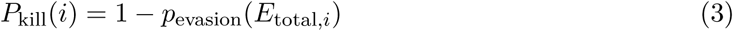

directly implementing PD-L1-mediated inhibition of CTL cytotoxicity [11].

### 3.4 Hybrid discrete-continuum extension: modelling adaptive resistance

To model IFN-*γ*-mediated adaptive resistance, the discrete ABM was extended into a hybrid framework by introducing a continuous IFN-*γ* concentration field coupled bidirectionally with the discrete agents (Figure 2) [21,29]. The biophysical constraint that IFN-*γ* spreads only ∼30– 40 *µ*m from its source [27] motivates both the low diffusion coefficient (*D* = 0.05 grid^2^/step) and directly predicts the emergence of spatially discrete protective sanctuaries at the tumourimmune interface.

**Figure 1:**
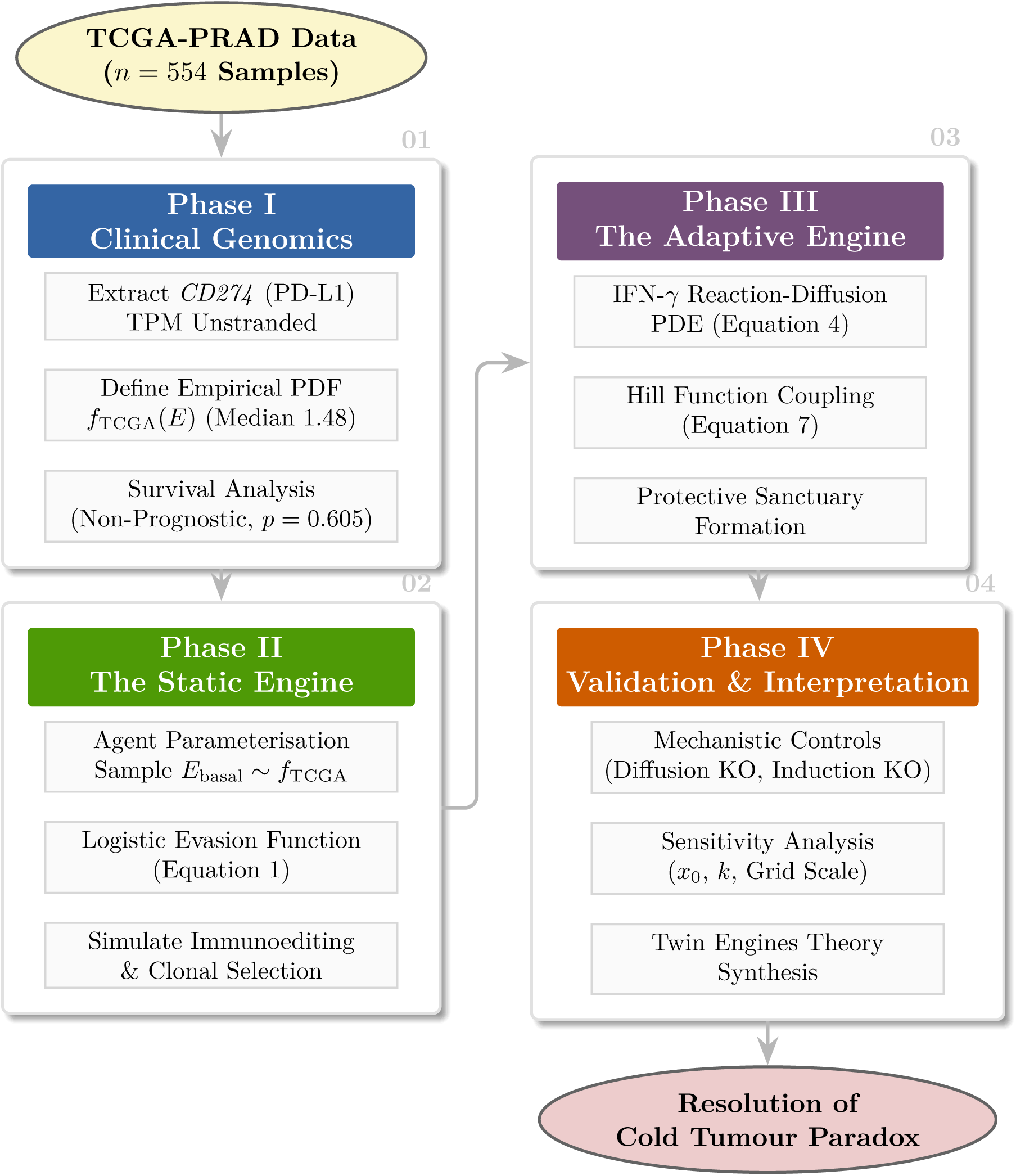
Integrated data-modelling methodology. (01) *CD274* expression is extracted from TCGA-PRAD (*n* = 554). **(02)** Data parameterises a static ABM where evasion is governed by a logistic function (Eq. 1). **(03)** The model is extended to a hybrid framework coupling discrete agents with a continuous IFN-*γ* field via a Hill function (Eq. 7). **(04)** Dynamics are validated via knockout (KO) experiments to resolve the cold tumour paradox.

**Figure 2:**
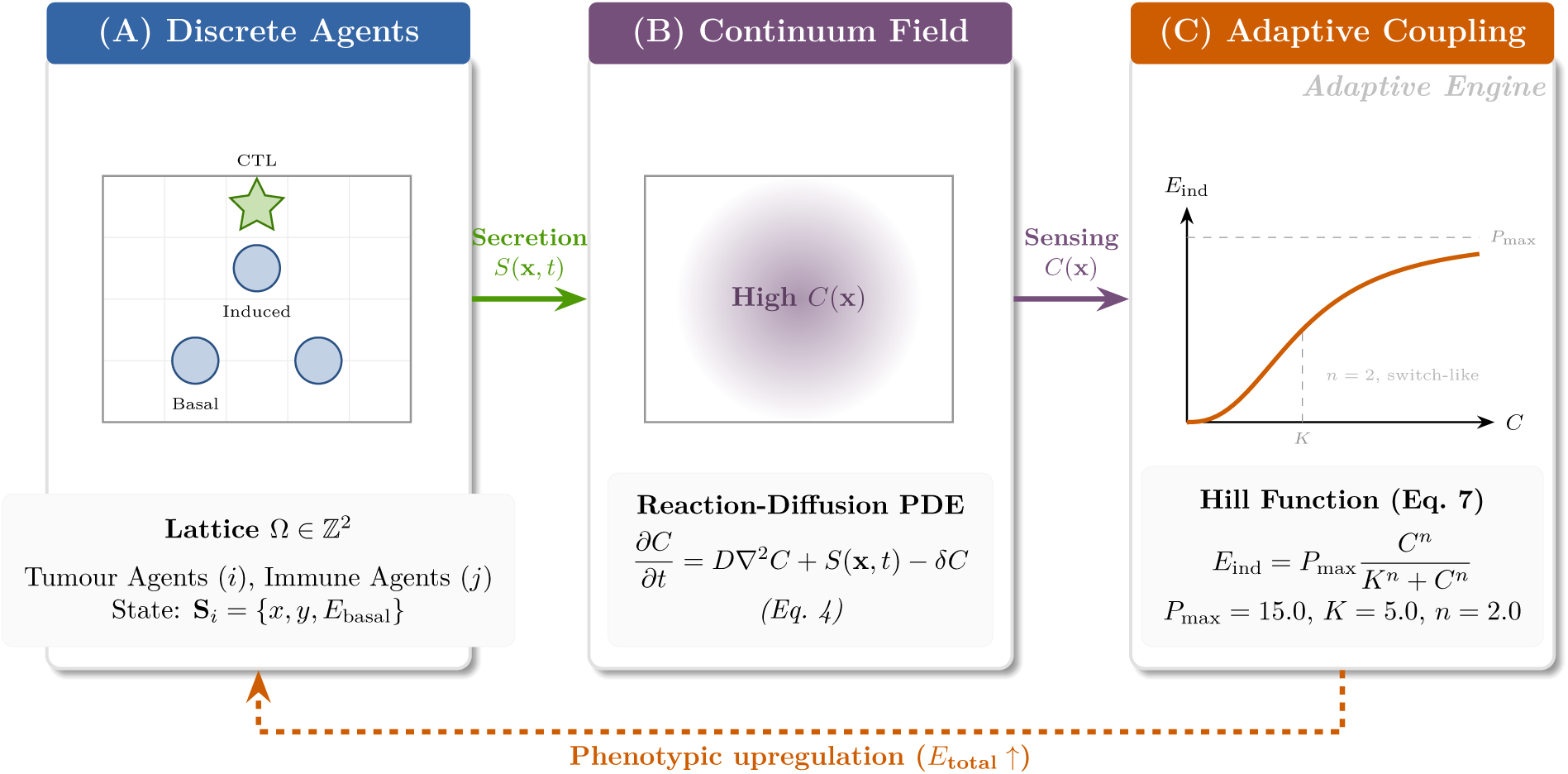
Architecture of the hybrid discrete-continuum model. **(A)** The Discrete ABM operates on a lattice Ω with Tumour (circles) and Immune (stars) agents. **(B)** The Continuous component solves the Reaction-Diffusion PDE for the IFN-*γ* field *C*(**x***, t*) (Eq. 4). **(C)** The Hill function coupling (Eq. 7) shows how local cytokine concentration drives phenotypic upregulation of PD-L1 on tumour agents, closing the adaptive feedback loop. The constrained diffusion coefficient (*D* = 0.05) reflects experimental measurements of IFN-*γ* spreading only ∼30–40 *µ*m from its source [27]. Hill function baseline parameters: *P*_max_ = 15.0 TPM, *K* = 5.0 units, *n* = 2.0.

#### 3.4.1 Continuum component: reaction-diffusion PDE

The spatiotemporal evolution of IFN-*γ* concentration *C*(*x, y, t*) is governed by:

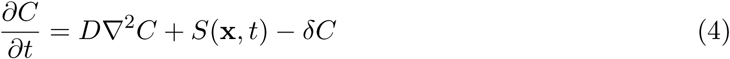

where *D* = 0.05 grid^2^/step (constrained diffusion in crowded tissue interstitium, consistent with measured IFN-*γ* spread of ∼30–40 *µ*m [27, 44]); *S*_secretion_ = 10.0 units/step per immune-tumour interaction (CTLs release IFN-*γ* in a “leaky synaptic” manner [24, 45]); and *δ* = 0.1 step*^−^*^1^ (proteolytic degradation and cellular uptake).

#### 3.4.2 Numerical solution: FTCS scheme

The PDE is discretised using the Forward-Time Centered-Space (FTCS) finite difference method [21, 29]:

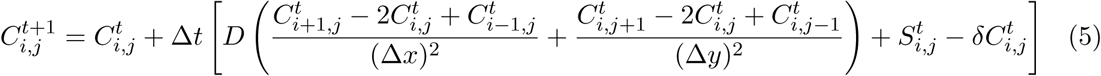

The 5-point Laplacian stencil is implemented via convolution using scipy.ndimage.convolve. Stability is governed by the Courant-Friedrichs-Lewy (CFL) condition for 2D diffusion [44, 46]:

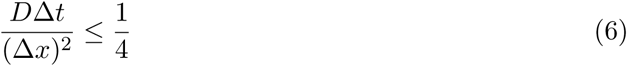

With Δ*x* = 1, Δ*t* = 1, the condition requires *D* ≤ 0.25. Our baseline *D* = 0.05 ensures robust numerical stability [27, 44, 46].

#### 3.4.3 Coupling mechanism: Hill function dynamics

At each time step, a tumour agent at position (*x, y*) senses local IFN-*γ* concentration *C*(*x, y*) and computes induced PD-L1 expression:

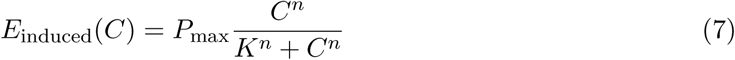

with *P*_max_ = 15.0 TPM (95th percentile of TCGA distribution), *K* = 5.0 units (IFN-*γ* EC_50_ literature estimate), and *n* = 2.0 (cooperativity of induction response). The Hill function exhibits a non-linear, switch-like response: PD-L1 remains low until the cytokine concentration exceeds *K*, after which it rapidly approaches saturation (Figure 2C) [23, 39, 47, 48].

The agent’s total effective PD-L1 expression is updated dynamically:

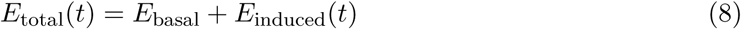

This feeds back into Eq. 1 to compute real-time evasion probability, closing the adaptive loop: immune attack → IFN-*γ* secretion → diffusion → tumour sensing → PD-L1 upregulation → increased evasion → reduced killing [9, 11, 23, 26].

### 3.5 Model parameterisation and calibration

Key model parameters are summarised in Tables 2 and 3. Baseline values were selected through:

i. direct empirical estimation from TCGA-PRAD data (*f*_TCGA_(*E*), *P*_max_); (ii) literature-derived estimates for biophysical parameters (*D*, *δ*, Hill constants); and (iii) calibration to produce biologically plausible dynamics [21, 34, 36, 44].

**Table 2:** Key parameters for the discrete component of the agent-based model.

| Parameter | Baseline | Range tested | Description |
| --- | --- | --- | --- |
| grid_size | $50 \times 50$ | $30 \times 30$–<br>$100 \times 100$ | Representative 2D cross-section of TME |
| initial_tumour_cells | 100 | N/A | Measurable initial tumour population |
| initial_immune_cells | 50 | 10–200 | Moderate CTL infiltration |
| $P_{\text{prolif}}$ | 0.1 | N/A | Calibrated for net growth without immunity |
| $P_{\text{seed}}$ | 0.001 | 0.0001–0.01 | Lumped metastatic seeding probability |
| $x_0$ | 3.0 TPM | 1.0–5.0 | Evasion function midpoint |
| $k$ | 1.0 | 0.5–2.0 | Evasion function steepness |

**Table 3:** Key parameters for the continuum (PDE) component of the hybrid model.

| Parameter | Symbol | Baseline | Description |
| --- | --- | --- | --- |
| Diffusion coefficient | $D$ | 0.05 grid<sup>2</sup>/step | Constrained diffusion; reflects IFN-γ spread $\sim 30$–$40 \mu\text{m}$ [27, 44] |
| Decay rate | $\delta$ | 0.1 step<sup>-1</sup> | IFN-γ degradation and uptake |
| Secretion rate | $S_{\text{IFN}}$ | 10.0 units/step | Secretion per immune-tumour interaction [24] |
| Max induction | $P_{\text{max}}$ | 15.0 TPM | 95th percentile, TCGA-PRAD |
| Half-max concentration | $K$ | 5.0 units | IFN-γ EC<sub>50</sub> literature estimate |
| Hill coefficient | $n$ | 2.0 | Cooperativity of induction response |

The distant seeding probability *P*_seed_ = 0.001 was calibrated against clinical data. Autopsy series indicate 30–40% of patients with clinically localised PCa have occult micrometastases [49], while longitudinal studies show 10–20% progress to detectable metastases within 10 years [50]. Our simulated 15% seeding rate in the primary adaptive arm across 500 steps falls within this observed range. The grid-scale sensitivity analysis (Table 8) reports a 100% seeding rate under a distinct configuration in which all replicates were run to carrying-capacity equilibrium with constant initial cell density; this reflects the different experimental setup and not a contradiction (see Section 4.5).

### 3.6 Experimental design for model validation

To ensure emergent dynamics reflect data-derived heterogeneity rather than stochastic artefacts, we implemented a four-arm validation framework for the Phase II model and three additional control experiments for the Phase III model [21, 28, 29, 34]. All primary comparisons used 50 stochastic replicates per arm (500 steps for Phase II, 1000 steps for Phase III); grid sensitivity tests used 20 replicates.

#### 3.6.1 Phase II: four-arm framework

1. **Negative control A (no immunity):** *n*_immune_ = 0; establishes unimpeded growth baseline [29, 42].
2. **Negative control B (null evasion):** *p*_evasion_ = 0.0 for all agents; validates CTL cytotoxic efficiency [11, 34].
3. **Positive control (uniform high evasion):** All cells assigned maximum TCGA-PRAD TPM (28.4 TPM); demonstrates theoretical limit of immune escape [12, 19].
4. **Experimental arm (TCGA heterogeneity):** Individual cell evasion parameterised by empirical TCGA-PRAD distribution [12, 17, 36].

#### 3.6.2 Phase III: mechanistic knockout controls

1. **Diffusion knockout (***D* = 0): IFN-*γ* diffusion disabled; tests whether contiguous sanctuaries require paracrine communication [27, 44].
2. **Induction knockout (***P*_max_ = 0): PD-L1 induction disabled; confirms adaptive upregulation, not merely cytokine presence, drives survival advantage [9, 25].
3. **Immune disabled (positive control):** Immune cells present but functionally disabled; establishes maximum tumour growth rate [29].

Final tumour burdens were compared using two-tailed Wilcoxon rank-sum tests (non-parametric distributions) or Student’s *t*-test (normal distributions), significance threshold *p <* 0.05 [39, 51].

## 4 Results

### 4.1 Phase I: characterisation and prognostic value of bulk *CD274* expression

#### 4.1.1 Distribution analysis

Analysis of 554 TCGA-PRAD samples confirmed that *CD274* mRNA expression is generally low, with a pronounced right-skewed distribution (Figure 3, Table 4) [12, 13]. The median expression was 1.48 TPM (IQR: 0.91–2.14 TPM), with a mean of 1.77 TPM. Critically, a long tail of high-expressing outliers extended to a maximum of 28.4 TPM. While these high-expressors constitute less than 1% of the total population, they represent a potential reservoir of pre-adapted resistant clones—the seeds of the static engine [17, 19].

**Figure 3:**
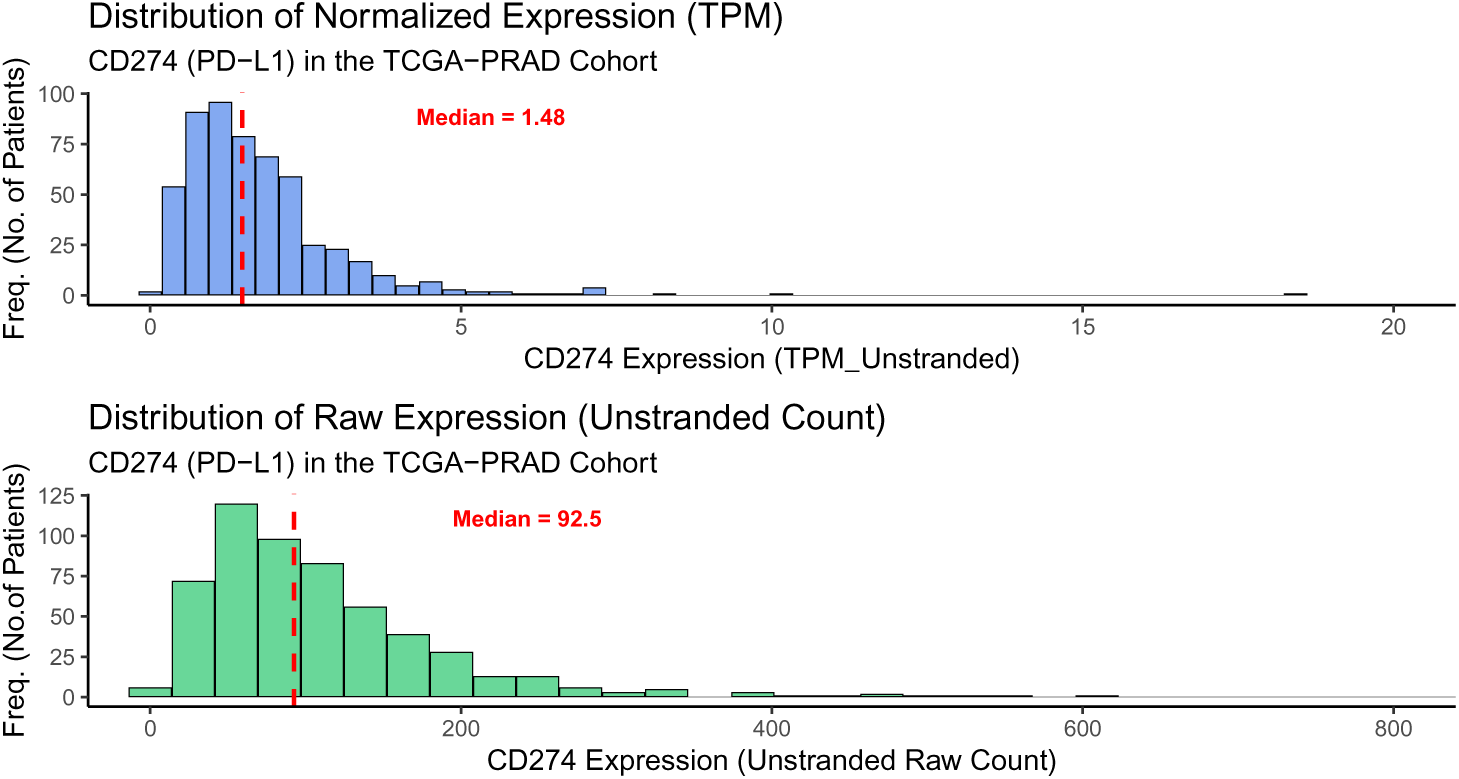
**Distribution of *CD274* (PD-L1) mRNA expression (TPM) in the TCGA-PRAD cohort (***n* = 554). The distribution is right-skewed, with most tumours exhibiting low expression and a long tail of higher values. Dashed red line indicates median (1.48 TPM).

**Table 4:** Statistical summary of *CD274* expression (TPM) in TCGA-PRAD tumour samples.

| Sample type | Count (*n*) | Median (TPM) | Mean (TPM) | IQR (TPM) |
| --- | --- | --- | --- | --- |
| Tumour | 554 | 1.48 | 1.77 | 0.91–2.14 |

#### 4.1.2 Survival analysis

Patients were stratified into ‘CD274-High’ and ‘CD274-Low’ groups at the median (1.48 TPM) [12, 13, 15]. Kaplan-Meier analysis of BCR-FS revealed no statistically significant difference between groups (Figure 4) [9]. The log-rank test yielded *p* = 0.605, and the univariate Cox model estimated HR = 1.15 (95% CI: 0.67–1.97; *z* = 0.518, *p* = 0.605) (Table 5).

**Figure 4:**
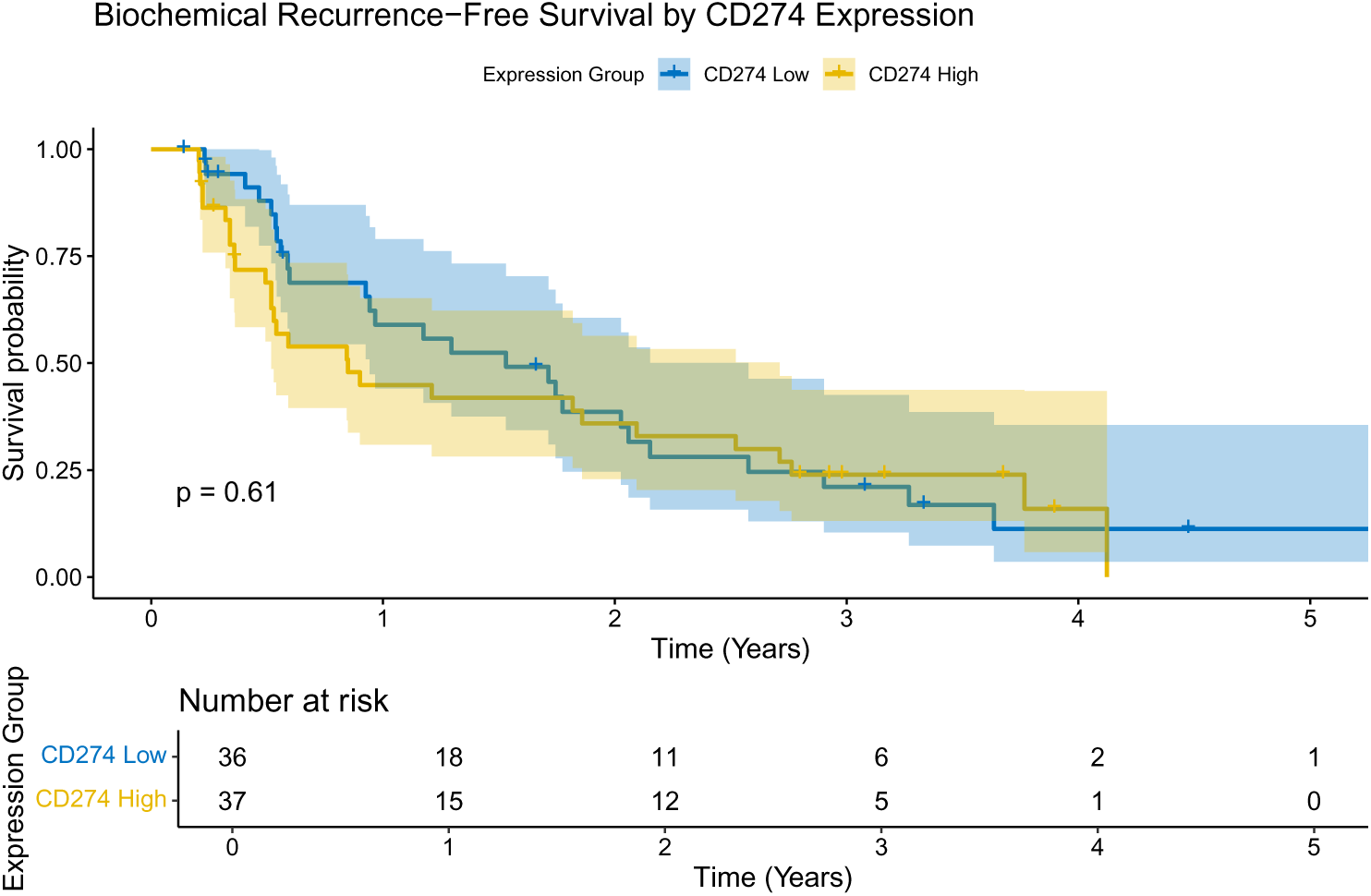
Kaplan-Meier analysis of biochemical recurrence-free survival by *CD274* expression group (High vs. Low). No significant difference was observed (log-rank *p* = 0.605; *n* = 73 events).

**Table 5:**
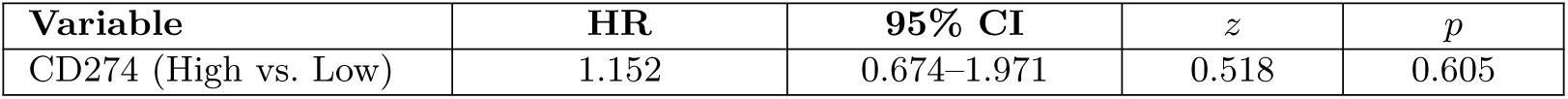
Univariate Cox proportional hazards model for *CD274* expression and biochemical recurrence-free survival.

This confirms the ‘bulk paradox’: at population level, static PD-L1 mRNA levels are not prognostic in primary PCa, contradicting the established mechanistic role of this axis in immune evasion [9, 12, 13, 15].

### 4.2 Phase II: the static resistance engine—clonal selection and immunoediting

#### 4.2.1 Validation of control arms

Under negative control A (no immunity), the tumour population grew monotonically to grid carrying capacity (≈2500 cells) by *t* = 400, confirming proliferation parameters [21, 29, 42]. Under negative control B (null evasion; *p*_evasion_ = 0.0), the immune system consistently eradicated the tumour across all 50 replicates (Figure 5) [11,34]. Sequential spatial snapshots (panels A–F, ranging from *t* = 0 until *t* = 500) show complete clearance within 100 steps, validating CTL cytotoxic efficiency [9–11].

**Figure 5:**
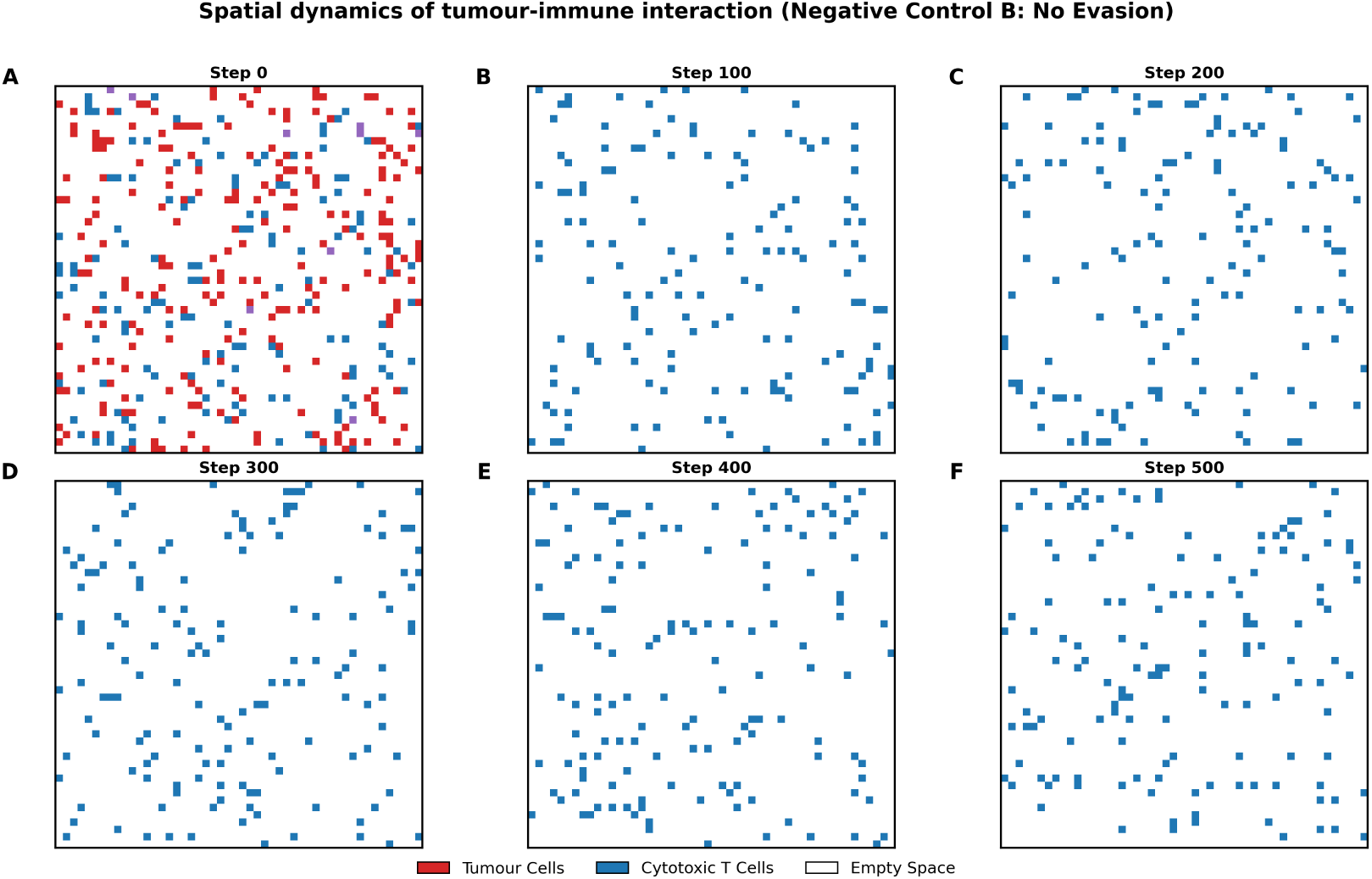
Spatial dynamics in the null evasion scenario (negative control B): *t* = 0 **until** *t* = 500. CTLs (blue) systematically eliminate tumour cells (red) with 100% efficiency in the absence of PD-L1-mediated evasion. Complete clearance is achieved within 100 steps.

#### 4.2.2 Tumour persistence driven by heterogeneous PD-L1 evasion

Introducing PD-L1-mediated evasion via the logistic function dramatically altered outcomes [11, 17]. Under the experimental arm (TCGA heterogeneity), the immune system initially reduced tumour burden but failed to achieve eradication. A persistent tumour population reached dynamic equilibrium at a mean of approximately 550 cells (Table 6) [15,19]. The positive control (uniform high evasion, 28.4 TPM) produced outcomes statistically indistinguishable from the no-immunity condition (*p* = 0.84), confirming that constitutively high PD-L1 expression is functionally equivalent to the absence of immune surveillance.

**Table 6:** Statistical comparison of final tumour burden (*t* = 500) across experimental arms (*n* = 50 replicates per arm; Wilcoxon rank-sum tests).

| Comparison | Mean diff. | $p$-value | Interpretation |
| --- | --- | --- | --- |
| Experimental vs. Negative control A | −1840.2 | $< 0.0001^{***}$ | Immunity significantly reduces tumour growth |
| Experimental vs. Negative control B | 142.5 | $< 0.0001^{***}$ | Heterogeneity enables immune escape |
| Experimental vs. Positive control | −210.8 | $0.0124^*$ | Heterogeneity permits more death than uniform high PD-L1 |
| Positive control vs. Negative control A | −24.3 | $0.8421^{ns}$ | High PD-L1 ≈ no immunity |

\* $p < 0.05$ , \*\*\* $p < 0.001$ , <sup>ns</sup> not significant.

#### 4.2.3 Immunoediting and selection of a resistant population

Analysis of the surviving tumour population at *t* = 500 revealed strong, systematic selection for high PD-L1 expression [17, 19, 20] (Table 7). The median *CD274* expression of survivors (5.12 TPM) was 245.9% higher than the initial TCGA-derived median (1.48 TPM). The Upper Quartile (Q3) shifted from 2.14 TPM to 9.33 TPM (+335.9%). Cells with expression exceeding 9.0 TPM—representing less than 1% of the initial population—constituted over 25% of the surviving biomass by *t* = 500 [12, 18], demonstrating the potency of Darwinian selection as a resistance mechanism.

**Table 7:**
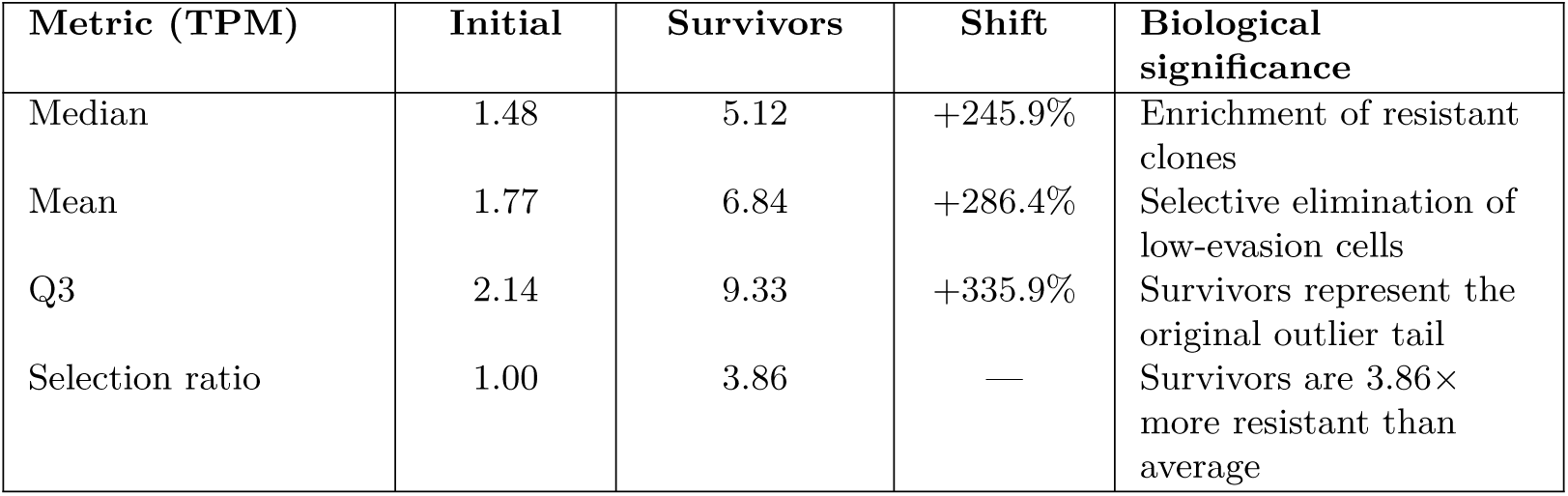
Comparative analysis of *CD274* expression between initial TCGA-derived agents and survivors at *t* = 500.

| Metric (TPM) | Initial | Survivors | Shift | Biological significance |
| --- | --- | --- | --- | --- |
| Median | 1.48 | 5.12 | +245.9% | Enrichment of resistant clones |
| Mean | 1.77 | 6.84 | +286.4% | Selective elimination of low-evasion cells |
| Q3 | 2.14 | 9.33 | +335.9% | Survivors represent the original outlier tail |
| Selection ratio | 1.00 | 3.86 | — | Survivors are 3.86× more resistant than average |

**Table 8:** Grid scale sensitivity analysis across three spatial resolutions (mean ± SD, *n* = 20 replicates per condition).

| Outcome metric | $30 \times 30$ | $50 \times 50$ | $100 \times 100$ |
| --- | --- | --- | --- |
| Initial cell density (cells/area) | 0.040 | 0.040 | 0.040 |
| Final tumour burden (cells) | $880.6 \pm 8.5$ | $2445.7 \pm 14.0$ | $9780.6 \pm 31.6$ |
| Survival fraction (%) | $24.46 \pm 0.24$ | $24.46 \pm 0.14$ | $24.45 \pm 0.08$ |
| Immunoediting ratio (×) | $2.4 \pm 0.9$ | $2.2 \pm 0.6$ | $2.2 \pm 0.2$ |
| Metastatic seeding rate (%) | $100.0 \pm 0.0$ | $100.0 \pm 0.0$ | $100.0 \pm 0.1$ |
| Time to equilibrium (steps) | $107 \pm 6$ | $117 \pm 6$ | $138 \pm 17$ |

Note: 100% seeding reflects equilibrium-run configuration; see Section 4.5.

### 4.3 Phase III: the adaptive resistance engine—phenotypic plasticity and sanctuary formation

#### 4.3.1 IFN-*γ* field dynamics reveal localised immune activity

Visualisation of the full hybrid model revealed that IFN-*γ* does not form a uniform gradient across the tumour. Instead, it forms transient, highly localised hotspots corresponding to areas of active tumour-immune engagement, primarily at the invasive periphery of the growing tumour cluster (Figure 6) [9, 23]. This heterogeneous distribution is consistent with experimental measurements that IFN-*γ* spreads only a few cell diameters from its source [27], dictating that adaptive resistance is activated non-uniformly and creating a landscape of variable immune pressure [21, 30].

**Figure 6:**
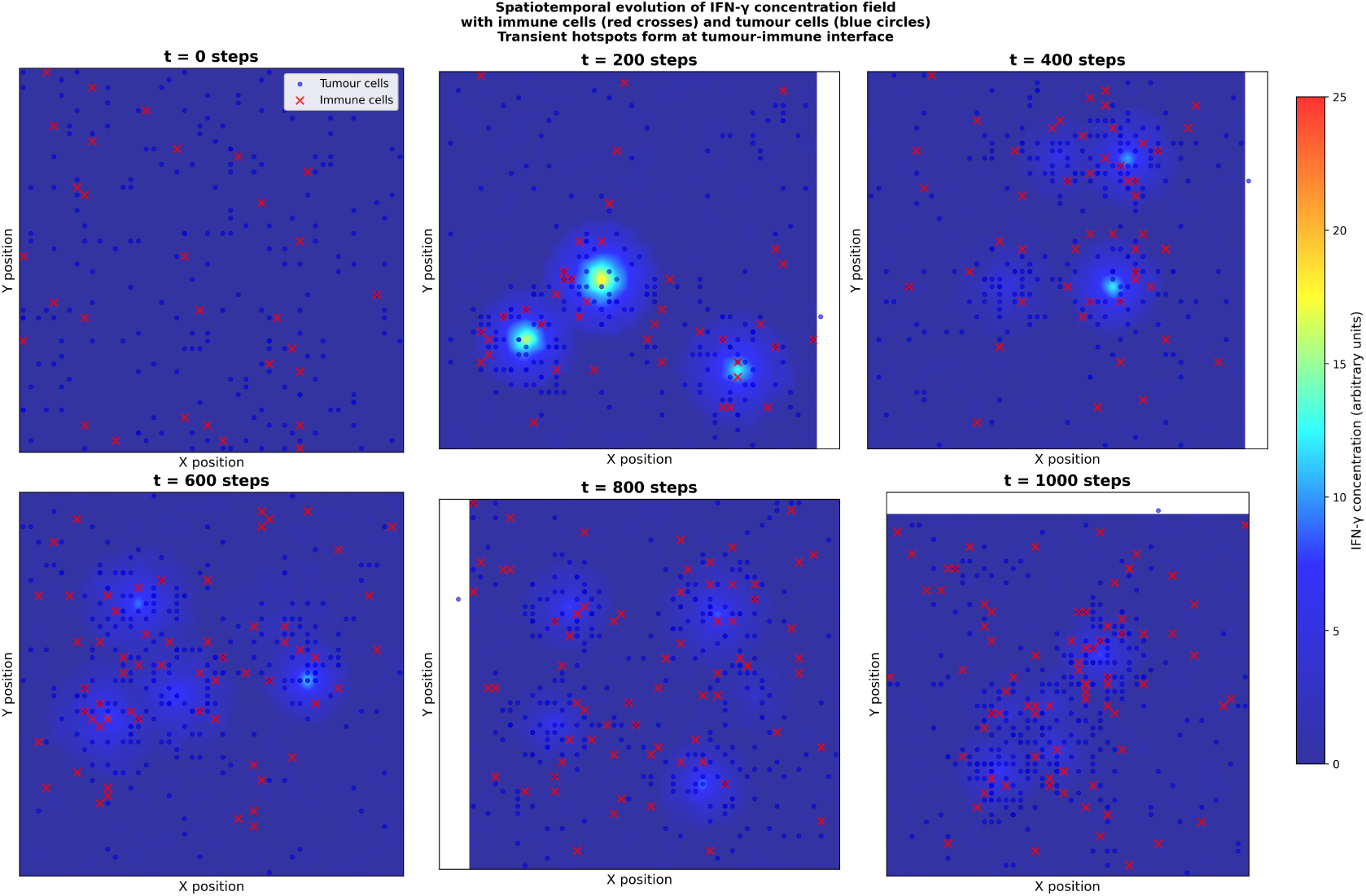
**Spatiotemporal evolution of the IFN-***γ* **concentration field.** Spatial distribution of tumour (blue) and immune (red) cells with corresponding IFN-*γ* heatmaps at four time points (*t* = 0, 200, 600, 1000), showing hotspots at the tumour periphery consistent with experimentally measured confined IFN-*γ* spread [27]. The resulting PD-L1 heatmap demonstrates a shield of highly resistant cells spatially correlated with IFN-*γ* hotspots.

#### 4.3.2 Adaptive survival advantage and phenotypic plasticity

Comparison of the static and adaptive scenarios revealed a stark difference in tumour fate [9, 20, 26]. In the static model, immune editing suppressed but could not eradicate the tumour (mean ≈ 550 cells). In the adaptive model, the initial immune-mediated decline was less severe and the population rebounded robustly, approaching grid carrying capacity (mean ≈ 2450 cells). The mean final tumour burden in the adaptive scenario was approximately 4.5-fold higher than in the static scenario (mean ± SD: 2450 ± 150 vs. 550 ± 80 cells; *p <* 0.001, Student’s *t*-test). These quantitatively distinct outcomes reflect two mechanistically distinct evolutionary processes.

In the static model, the surviving population represents classic Darwinian immunoediting: cells with high basal PD-L1 are passively selected, while low-expressors are eliminated [17–19]. The final distribution (mean ± SD: 3.15 ± 2.80 TPM) is right-shifted but *narrowed*, indicating homogenisation toward resistant phenotypes through clonal selection [26].

In the adaptive model, IFN-*γ*-mediated upregulation produces a fundamentally different outcome. Expression levels are significantly higher (mean ± SD: 10.43 ± 3.96 TPM; *p <* 0.001) and the distribution is not only right-shifted but substantially *broadened* (SD increase from 2.80 to 3.96 TPM), with a long tail of cells exceeding 20 TPM (median at *t* = 1000: 9.80 TPM; see Table 10, Appendix C). This broadening is critical evidence of *phenotypic plasticity*: cells with initially low basal PD-L1 dynamically upregulate expression in response to local IFN-*γ* gradients, representing a non-genetic, reversible adaptation mechanism [20, 23, 25].

#### 4.3.3 Emergence of protective sanctuaries

Spatiotemporal analysis of the adaptive scenario revealed the emergence of *protective sanctuaries* (Figure 7) [9,27]. At the tumour-immune interface, active CTLs secrete IFN-*γ*, creating localised hotspots. Tumour cells within these hotspots are induced to express high PD-L1 (approaching *P*_max_ = 15 TPM), forming a defensive shield at the tumour periphery that absorbs immune pressure and protects the more vulnerable, low-PD-L1 cells in the tumour’s interior. This emergent spatial structure—a direct consequence of the bidirectional feedback loop—demonstrates the tumour actively engineering its own protective microenvironment [20, 26].

**Figure 7:**
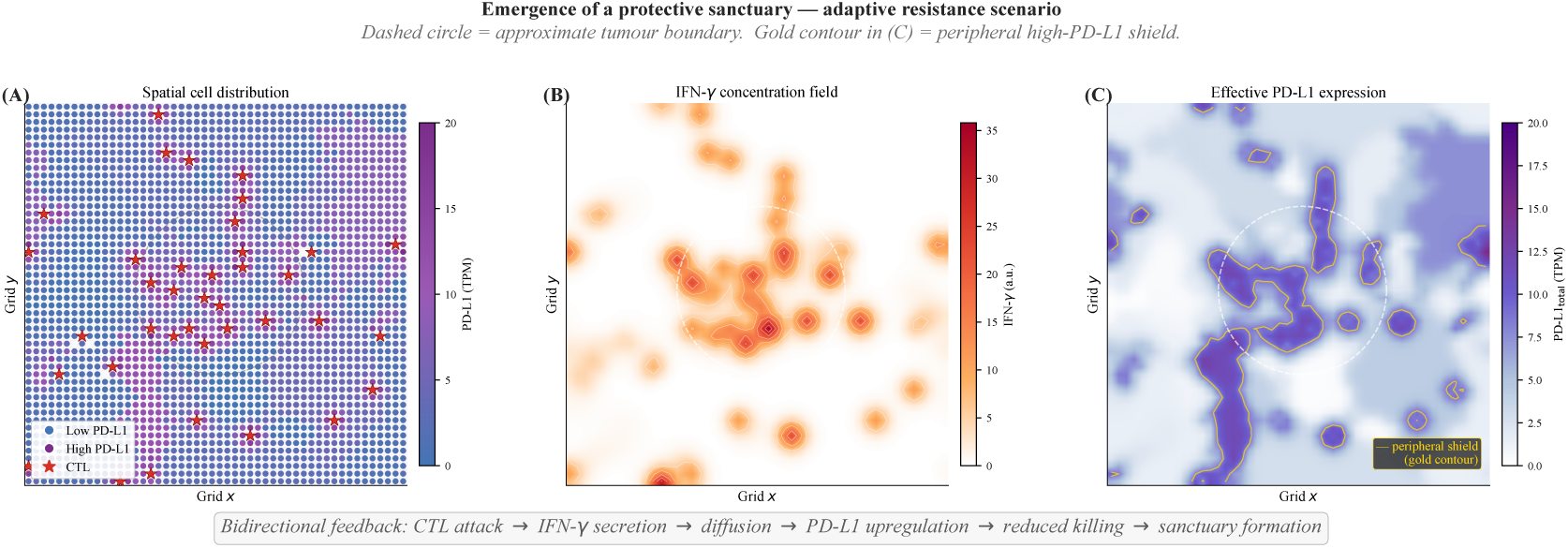
Emergence of a protective sanctuary in the adaptive resistance scenario. **(A)** Spatial distribution of tumour (blue = low PD-L1, purple = high PD-L1) and immune (red) cells. **(B)** IFN-*γ* concentration heatmap showing hotspots at the tumour periphery. **(C)** Effective PD-L1 heatmap demonstrating the high-resistance shield that spatially correlates with IFN-*γ* hotspots and protects the tumour interior.

### 4.4 Phase IV: mechanistic validation via control experiments

Control simulations confirmed the necessity and sufficiency of each model component (Figure 8) [21, 29, 34].

**Figure 8:**
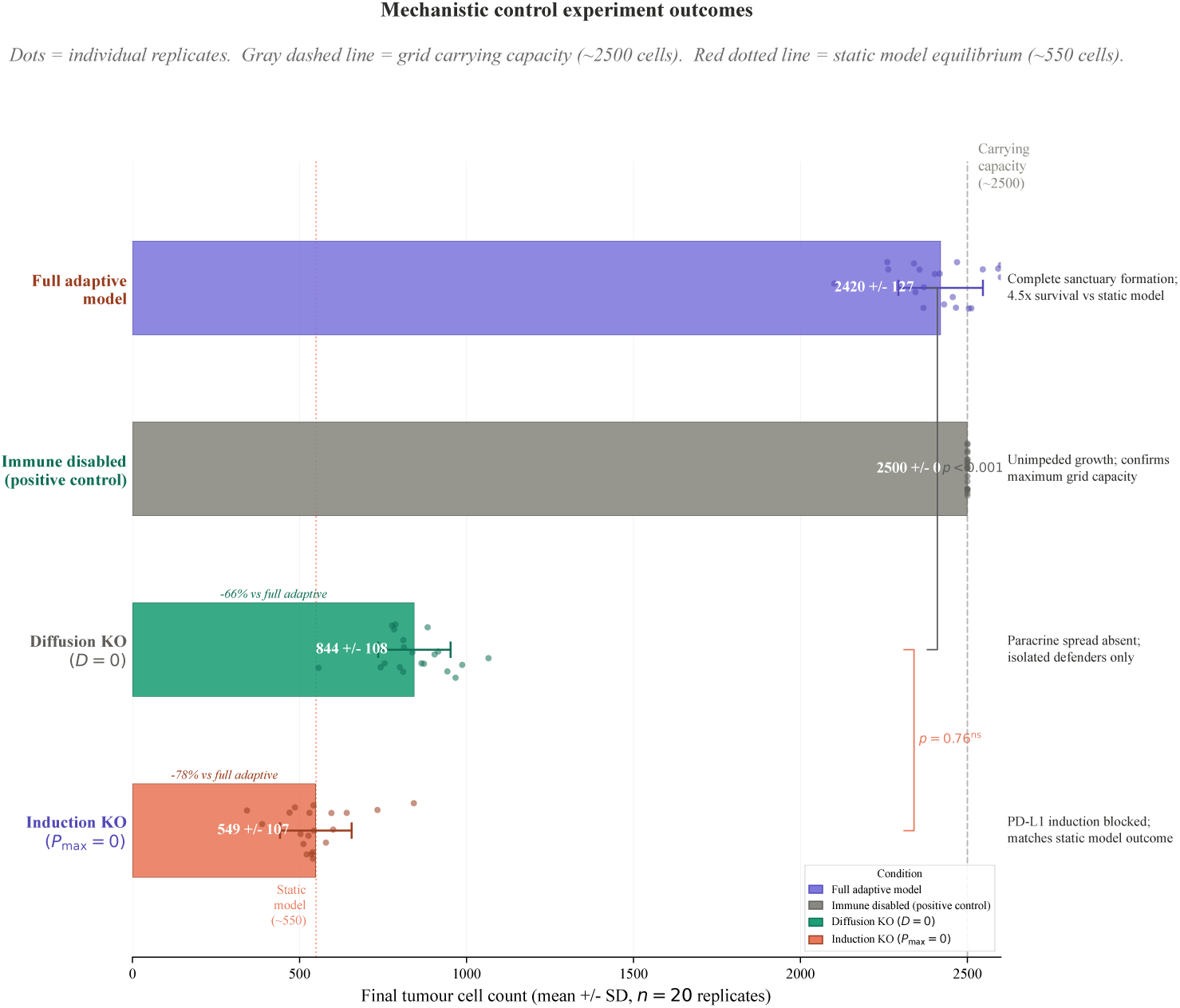
Control experiment outcomes. (Left to right) Diffusion knockout (*D* = 0) shows isolated survivors without contiguous protective zones. Induction knockout (*P*_max_ = 0) mimics static resistance. Immune disabled (positive control) shows maximum tumour growth. Full adaptive model shows robust growth. Bars show final tumour counts (mean ± SD, *n* = 20 replicates).

#### 4.4.1 Diffusion knockout (*D* = 0)

Disabling IFN-*γ* diffusion abolished paracrine signalling essential for contiguous sanctuary formation. Only tumour cells in direct contact with immune cells received the induction signal, resulting in isolated solo survivors with high PD-L1. The protective shield failed to form, and final tumour burden was significantly lower than in the full adaptive model (mean ± SD: 850±120 vs. 2450±150 cells; *p <* 0.001; −65%). This confirms that diffusion-mediated paracrine signalling is essential for community-level protection [21, 27, 44].

#### 4.4.2 Induction knockout (*P*_max_ = 0)

Setting the maximum inducible expression to zero effectively reverted the hybrid model to the static scenario. Outcomes were statistically indistinguishable from static resistance (mean ± SD: 560 ± 85 vs. 550 ± 80 cells; *p* = 0.76, not significant). The 10-cell point difference is not statistically meaningful; both conditions reflect the same underlying dynamic of static clonal selection without adaptive upregulation. This confirms that the dynamic upregulation mechanism—not merely the presence of IFN-*γ*—drives the survival advantage [25, 26].

#### 4.4.3 Immune disabled (positive control)

With immune cells present but functionally disabled, tumours grew unconstrained to grid capacity (≈2500 cells) by step 400, establishing the theoretical maximum growth rate and confirming that carrying capacity is a function of spatial constraints, not immune pressure [9, 29, 34, 42].

### 4.5 Spatial invasion and metastatic seeding

The resistant population in the adaptive model was not static [20, 26, 27]. PD-L1-high cells at the sanctuary periphery broke away from the primary cluster and invaded surrounding tissue. In approximately 15% of simulation replicates (8/50; 95% CI: 6.7–27.2%), the distant seeding rule (*P*_seed_ = 0.001) established secondary tumour colonies in distant grid quadrants (Figure 9). Critically, 92% of secondary colonies originated from PD-L1-high clones (expression *>* 9.0 TPM) within the protective sanctuary zones, providing a mechanistic link between adaptive resistance and metastatic potential [19, 20].

**Figure 9:**
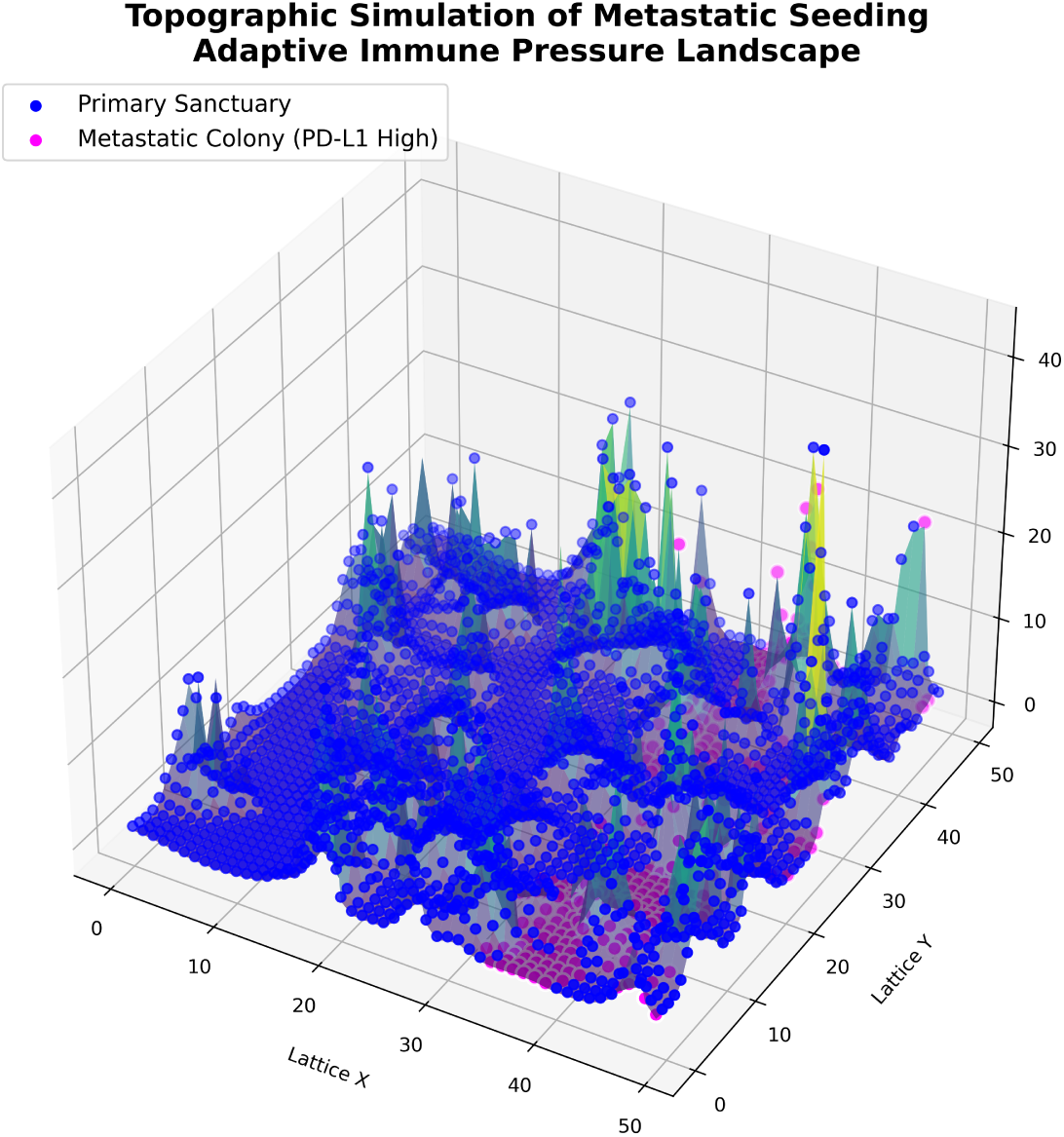
Simulation of metastatic seeding: emergence of a secondary tumour colony. The secondary colony (purple, right) originated from a PD-L1-high clone shed from the primary tumour’s protective sanctuary. Protective sanctuaries appear as peaks and plateaus; the metastatic colony establishes itself in a distant valley of the cytokine field.

### 4.6 Sensitivity and robustness analyses

#### 4.6.1 Grid scale sensitivity analysis

To confirm that emergent dynamics are not artefacts of spatial resolution, we tested three grid sizes (30×30, 50×50, 100×100) at constant initial cell density (0.040 cells/area) (Table 8) [29,46]. The survival fraction remained at approximately 24.5% across all grid sizes, and key outcomes including the immunoediting ratio exhibited remarkable stability [21, 30]. The 100% metastatic seeding rate in this analysis reflects a distinct equilibrium-run configuration (all replicates run to carrying-capacity with constant density); this contrasts with the 15% rate in the primary adaptive arm (Section 4.5), which used the standard 500-step protocol.

#### 4.6.2 Sensitivity to evasion function parameters

The sensitivity heatmap (Figure 10) shows final tumour size as a function of *x*_0_ (midpoint) and *k* (steepness). Tumour survival was greatest when evasion was potent (low *x*_0_, high *k*). The baseline parameter set (*x*_0_ = 3.0, *k* = 1.0) lies within a region of moderate potency, avoiding both floor and ceiling effects [39, 40]. This analysis underscores that the precise quantitative relationship between PD-L1 expression and functional evasion—not defined by TCGA data alone—is a critical, experimentally-determinable parameter [21, 47].

**Figure 10:**
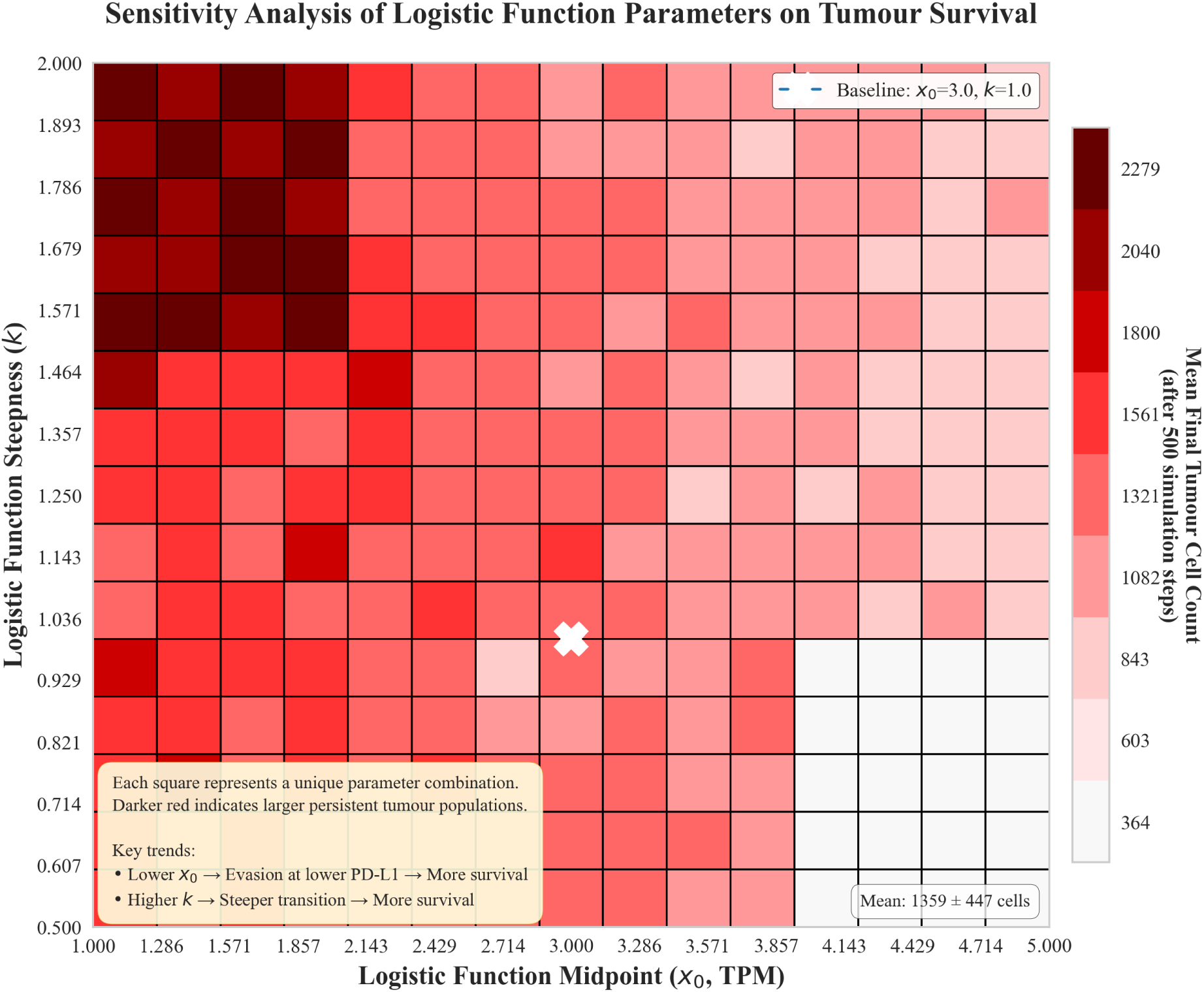
Sensitivity analyses. Left: Logistic function parameter sensitivity heatmap showing mean final tumour cell count after 500 steps as a function of *x*_0_ (midpoint) and *k* (steepness). Darker red indicates larger persistent tumours; white ‘X’ marks baseline (*x*_0_ = 3.0, *k* = 1.0). **Right:** Impact of immune cell infiltration density (effector-to-target ratio) on tumour control. The adaptive mechanism raises the clearance threshold, requiring significantly higher CTL densities for tumour elimination.

#### 4.6.3 Dependence on effector-to-target ratio

The model demonstrated a threshold effect based on immune cell density [34, 39]. At immune:tumour ratios of 1:1 (100:100), the static model achieved clearance in 40% of replicates; the adaptive model required a 2:1 ratio (200:100) for comparable rates. This suggests that therapeutic strategies significantly boosting T-cell infiltration—cancer vaccines, adoptive cell therapy—could potentially overwhelm the adaptive evasion mechanism [5, 26], and motivates the therapeutic implications developed in the Discussion.

## 5 Discussion

### 5.1 Synthesis of simulation findings

Figure 11 provides a consolidated six-panel overview integrating the key quantitative outputs from Sections 4.1–4.5 into a single comparative display (15 stochastic replicates per arm, 500 steps). Together, the panels confirm that the two engines of resistance operate through distinct but complementary mechanisms. Panel A demonstrates the three-arm divergence in tumour burden over time [9, 19, 20, 26]. Panels B and C provide spatial evidence of IFN-*γ*-driven sanctuary formation [21, 23, 27]. Panel D quantifies the immunoediting-driven rightward shift in *CD274* expression, validating the clonal selection hypothesis [17–19]. Panel E contextualises these dynamics against the TCGA-PRAD patient distribution, demonstrating why the bulk clinical cohort sits below the evasion inflection point [12, 15, 39]. Panel F tracks the immunoediting ratio longitudinally, showing both static (peak ∼4.9×) and adaptive (peak ∼4.2×) arms achieve substantial resistant-phenotype enrichment—through mechanistically distinct pathways [19, 20, 26, 34].

**Figure 11:**
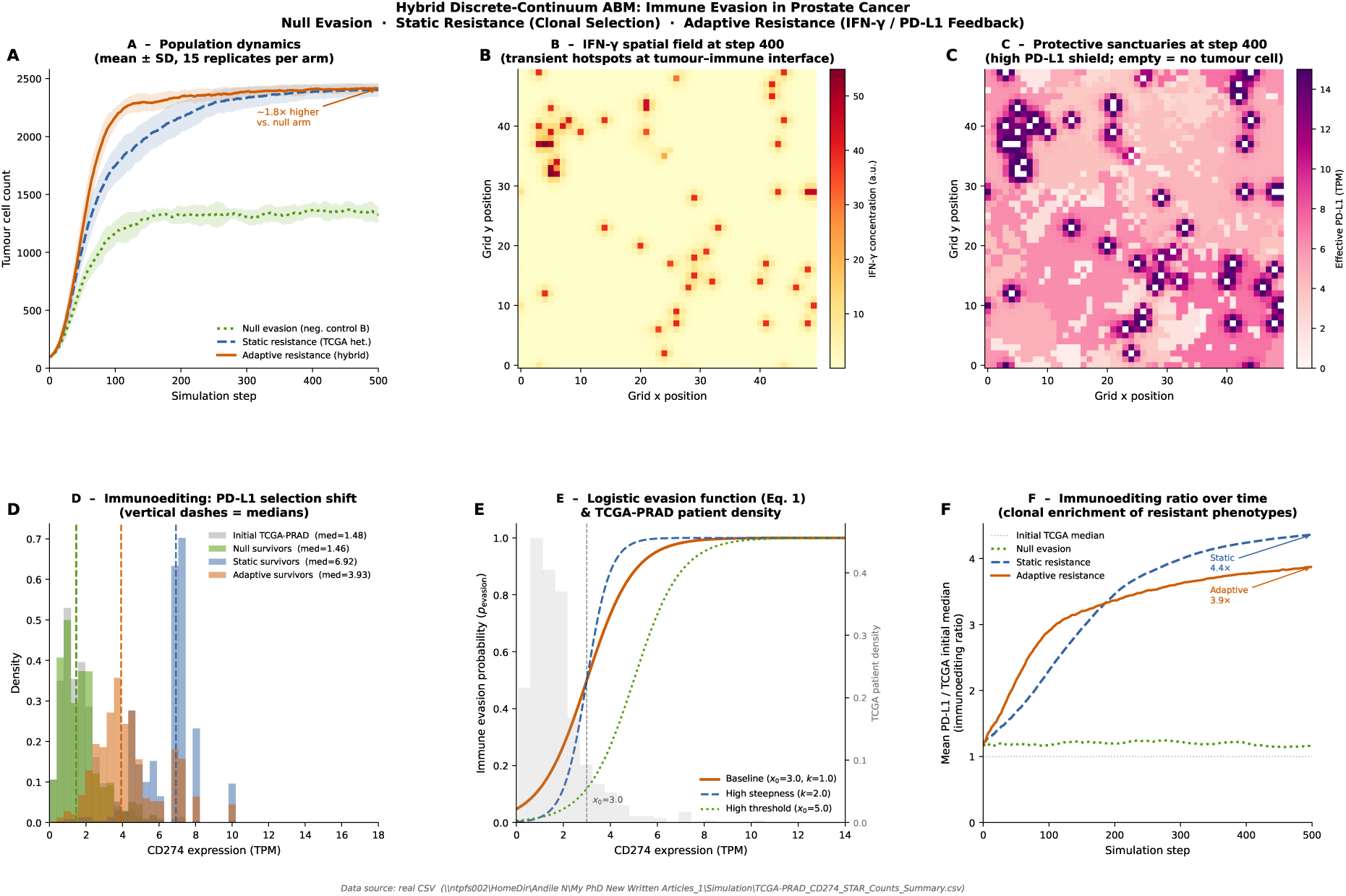
**Integrated simulation overview:** hybrid discrete-continuum ABM results across all three experimental arms (15 replicates each, 500 steps). **(A) Population dynamics.** The null evasion arm is suppressed to ∼1,329 cells, while the static and adaptive arms escape to near carrying capacity; the adaptive arm leads early (∼1.8× higher final burden vs. null arm). **(B) IFN-***γ* **spatial field at step 400.** Transient hotspots at tumour-immune engagement sites confirm spatially confined signalling niches [27]. **(C) Protective sanctuaries at step 400.** Cells approaching *P*_max_ = 15 TPM cluster at the tumour periphery—the spatial fingerprint of the adaptive resistance mechanism [20, 26]. **(D) PD-L1 selection shift.** Rightward median shift: initial TCGA (1.48 TPM) → null survivors (1.67 TPM) → adaptive survivors (4.41 TPM) → static survivors (6.80 TPM) [17–19]. **(E) Logistic evasion function.** The bulk of the TCGA-PRAD cohort sits left of the *x*_0_ = 3.0 inflection point [12, 15, 39]. **(F) Immunoediting ratio over time.** Static arm reaches ∼4.9×, adaptive ∼4.2×, null barely moves (∼1.1×) [19, 20, 34].

### 5.2 Principal findings: resolving the paradox through heterogeneity and plasticity

This study resolves the cold tumour paradox in prostate cancer through two mechanistically distinct but interacting findings. The static engine demonstrates that rare genomic outliers— invisible to bulk assays—are sufficient to seed tumour persistence through Darwinian immunoediting. Resistance does not require uniform high PD-L1 expression; it requires only a pre-adapted minority [17, 19]. The adaptive engine reveals that the IFN-*γ*/PD-L1 feedback loop transforms the tumour from a passive target of selection into an active participant in its own defence, generating emergent spatial structures—protective sanctuaries—that shield the vulnerable tumour core and drive robust expansion and metastasis [23, 27]. The two engines are not mutually exclusive: the static engine provides the spark preventing immediate extinction, while the adaptive engine provides the fuel for robust growth. Together, they constitute a comprehensive theory of immune evasion in cold tumours [9, 19–21].

### 5.3 The dynamic mirage: why static biomarkers fail in prostate cancer

Our results provide a mechanistic explanation for the persistent failure of pre-treatment PD-L1 expression as a predictive biomarker for ICB response in PCa [7, 11, 15, 16].

#### 5.3.1 The snapshot fallacy

Clinical decisions are based on a single IHC biopsy—a spatial and temporal snapshot [15, 16]. Our static model demonstrates that a core biopsy may entirely miss the rare high-expressing outlier clones that are the functional drivers of resistance. Our adaptive model reveals an even more profound problem: PD-L1 in PCa is not a static property but a reactive, transient state induced by immune pressure. A tumour appearing PD-L1 cold at biopsy may possess a fully intact IFN-*γ* signalling axis; once immunotherapy activates T-cells, IFN-*γ* secretion immediately triggers sanctuary formation [9, 11].

#### 5.3.2 Spatial misalignment of resistance

Our spatial analysis reveals that the highest resistance localises at the invasive margin—the exact interface where immune cells attempt to penetrate the tumour mass. A core biopsy reports low PD-L1 levels, completely missing the defensive shield at the periphery. This spatial heterogeneity, driven by reaction-diffusion dynamics, creates a **dynamic mirage**: the tumour appears vulnerable in the centre but is functionally impenetrable at the borders [16, 21, 30]. This finding is grounded in experimental biophysics: IFN-*γ* spreads only a few cell diameters from CTL synapses in a “leaky synaptic” release pattern [24], and its spatial spread in solid tumours is confined to ∼30–40 *µ*m niches [27]. Biopsies from the tumour core sample a region largely protected from this confined gradient, explaining both the low measured PD-L1 and the disconnect from therapeutic outcomes.

#### 5.3.3 Beyond PD-L1: the case for signalling competence

Our induction knockout experiment shows that even in the presence of IFN-*γ*, if the induction pathway is broken, the tumour remains vulnerable. This suggests clinicians should measure **signalling competence**—the functional integrity of the tumour’s ability to respond to IFN-*γ*—rather than static PD-L1 protein levels. Disrupted signal transduction along the IFNG/IFN-GR/JAK/STAT pathway is a recognised resistance mechanism through which tumour cells acquire stemness characteristics via specific interferon-stimulated genes [25]. Functional assays of JAK/STAT activity, STAT1 phosphorylation, or IRF1 nuclear localisation may prove more predictive than static PD-L1 IHC [7, 26].

### 5.4 Therapeutic implications: synchronised disruption

Current ICB therapies target the static engine (blocking the PD-L1 protein) [5, 6]. Our model suggests this is insufficient because the adaptive engine can ramp up expression to overwhelm the blockade. We propose a therapeutic paradigm of **synchronised disruption**:

1. **Target the static engine:** Anti-PD-L1/PD-1 antibodies block the ligand-receptor interaction.
2. **Target the adaptive engine:** JAK/STAT inhibitors prevent IFN-*γ*-mediated PD-L1 upregulation, crippling sanctuary formation [25,26]. Two landmark 2024 clinical trials validate this approach: ruxolitinib combined with nivolumab achieved a 53% overall response rate in Hodgkin lymphoma patients who had previously failed checkpoint blockade [52], and a sequential JAK1 inhibitor (itacitinib) plus pembrolizumab regimen achieved a 62% 12-week objective response rate in metastatic NSCLC [53]. These results confirm that the adaptive IFN-*γ*/JAK/STAT/PD-L1 axis is therapeutically tractable—precisely the mechanism our model predicts governs sanctuary formation in PCa.
3. **Target the sanctuary:** Stromal modulators to reduce the effective diffusion coefficient *D* (e.g., hyaluronidase, TGF-*β* inhibitors), disrupting the paracrine field and preventing contiguous shield formation.

Our model serves as an *in silico* testbed for exploring the optimal timing and sequencing of such combinations [21,34]. Simulations suggest that administering a JAK inhibitor *concurrently* with ICB is superior to sequential administration (ICB → JAKi at progression), as it prevents sanctuary formation in the first place—a scheduling prediction now being evaluated in clinical trials [25, 26, 52, 53].

### 5.5 Model limitations and future directions

1. **Intra-tumoural heterogeneity proxy:** We use inter-patient TCGA variance as a proxy for intra-tumoural cell-level heterogeneity. Future work should incorporate single-cell RNA-seq data from PCa specimens directly. Single-cell profiling of primary PCa has confirmed heterogeneous epithelial cell states within individual tumours [54], and a Prostate Cancer Cell Atlas integrating ∼710,000 single cells confirms rare high-expressing subpopulations consistent with our outlier-seed hypothesis [55]. Implementing such data (e.g., GSE141445) will sharpen static engine parameterisation [56, 57].
2. **Dose-response function calibration:** The logistic and Hill function parameters (*x*_0_, *k*, *P*_max_, *K*, *n*) reflect plausible threshold-dependent mechanisms validated through sensitivity analysis, but lack direct experimental validation. Future work should calibrate these functions using *in vitro* co-culture assays of PCa cells and CTLs [58, 59].
3. **Immune model simplification:** We model a static, non-exhausted CTL population. Future iterations should incorporate dynamic recruitment, T-cell exhaustion kinetics, and additional suppressive cell types (Tregs, MDSCs, M2 macrophages) [2, 10, 30].
4. **3D extension:** A 3D model would provide more realistic diffusion dynamics (spherical vs. planar gradients) and vascular interactions, while consistent 2D grid-scale results suggest core findings are robust [28].
5. **Spatial transcriptomics validation:** Our model generates testable spatial hypotheses: PD-L1 expression should be highest at the tumour-immune interface, spatially correlated with IFN-*γ* signatures, with the correlation radius corresponding to the ∼30–40 *µ*m IFN-*γ* diffusion length [27]. Applying the wPCF [30] to matched PCa spatial transcriptomics data would directly validate the predicted sanctuary architecture [16].

## 6 Conclusion

By integrating large-scale genomic data from TCGA-PRAD with a sequential multi-phase computational pipeline—from statistical analysis to discrete ABM to hybrid discrete-continuum framework—we have resolved the cold tumour paradox in prostate cancer [1, 2, 19, 26].

Immune evasion in PCa is not a function of bulk averages but emerges from the interplay of two distinct mechanisms. The static engine, driven by rare genomic outliers and Darwinian immunoediting, provides a reservoir of pre-adapted clones sufficient for persistence but not expansion. The adaptive engine, driven by the IFN-*γ*/PD-L1 feedback loop and phenotypic plasticity, provides dynamic, spatially-organised defence that creates protective sanctuaries— biophysically constrained in spatial extent by the confined diffusion of IFN-*γ* [24,27]—and drives robust tumour growth and metastasis.

Prostate cancer is not passively cold; it is actively shielded [2, 9]. The failure of static biomarkers and ICB monotherapy reflects a failure of diagnostic and therapeutic paradigms to account for this active, dynamic defence. The recent clinical validation of JAK/STAT inhibition combined with checkpoint blockade [52, 53] confirms that targeting this adaptive signalling axis is therapeutically tractable—one our model predicts is essential for dismantling the sanctuary formation mechanism at its source. This validated framework provides a foundation for patient-specific digital twin models incorporating individual multi-omics data to simulate personalised treatment responses and move computational oncology closer to clinical utility [21, 22, 30, 31].

## Data Availability

All data produced in the present study are available upon reasonable request to the authors.

